# Cross-sectional assessment of predictors for COVID-19 vaccine uptake: an online survey in Greece

**DOI:** 10.1101/2021.09.23.21264009

**Authors:** Petros Galanis, Irene Vraka, Olga Siskou, Olympia Konstantakopoulou, Aglaia Katsiroumpa, Ioannis Moisoglou, Daphne Kaitelidou

**Affiliations:** Clinical Epidemiology Laboratory, Faculty of Nursing, National and Kapodistrian University of Athens, Athens, Greece; Department of Radiology, P & A Kyriakou Children’s Hospital, Athens, Greece; Center for Health Services Management and Evaluation, Faculty of Nursing, National and Kapodistrian University of Athens, Athens, Greece; Pulmonary Clinic, General Hospital of Lamia, Lamia, Greece

**Keywords:** COVID-19, general population, vaccine uptake, Greece

## Abstract

**Background:** A high level of COVID-19 vaccine uptake in the general population is essential to control the pandemic.

**Objective:** To estimate the percentage of the general population vaccinated against the COVID-19 and to investigate the factors associated with COVID-19 vaccine uptake.

**Methods:** We conducted an online cross-sectional study in Greece during August 2021. We included individuals over 18 years of age. Independent variables included socio-demographic data of the participants and attitudes towards COVID-19 vaccination and pandemic. Our outcome variable was COVID-19 vaccination status, measured through “yes/no” answers.

**Results:** Most participants had been vaccinated against the COVID-19 (87.8%), while about half had been vaccinated against the influenza (52.5%). Multivariate analysis identified that increased age and higher level of education were associated with an increased likelihood of COVID-19 vaccination. Also, participants working in health services, participants without a previous COVID-19 diagnosis and those with previous seasonal influenza vaccination history had a greater probability to take a COVID-19 vaccine. Additionally, increased self-perceived severity of COVID-19, knowledge regarding COVID-19, and trust in COVID-19 vaccines and scientists were associated with COVID-19 vaccine uptake. On the other hand, the likelihood of vaccination was lower for participants who were more concerned about the side effects of COVID-19 vaccination.

**Conclusions:** Understanding the factors affecting individuals’ decision to take a COVID-19 vaccine is essential to improve the COVID-19 vaccination coverage rate. Policymakers and scientists should scale up their efforts to increase the COVID-19 vaccination rate among specific population groups such as young people, people with low level of education, people with negative attitudes towards vaccination, etc.

## Introduction

Until September 2021, the Coronavirus disease 2019 (COVID-19) pandemic has caused more than 4.7 million deaths and 230 million cases (Worldometer, 2021). COVID-19 vaccination is regarded as the most promising means of controlling the COVID-19 pandemic. However, there is a critical need for effective vaccine uptake in the general population to approach herd immunity. COVID-19 vaccine acceptance shows a great variability across countries or regions which may affect the efforts to contain the pandemic (Aw et al., 2021). According to meta-analyses (Snehota et al., 2021; Q Wang et al., 2021), the average COVID-19 vaccine acceptance rate in the general population is about 73% which is lower than the percentage estimated to be sufficient for reaching herd immunity threshold with a COVID-19 vaccine that is at least 80% effective (Bartsch et al., 2020). Moreover, the longer the COVID-19 pandemic lasts, the smaller rate of the general population wants to get vaccinated (Snehota et al., 2021). The situation may get worse as the real-world COVID-19 vaccine uptake may be lower than the acceptance rate.

Predictors of COVID-19 vaccine acceptance are well known. In particular, socio-demographic variables (e.g. gender, age, race/ethnicity, and educational level), influenza vaccination history, self-protection from COVID-19, risk perception of COVID-19 infection, fear of COVID-19, concerns about the efficacy, side effects and safety of COVID-19 vaccines, and trust in authorities are the most prominent factors associated with COVID-19 vaccination willingness (AlShurman et al., 2021; Joshi et al., 2021; Q Wang et al., 2021).

Unfortunately, the literature on predictors for COVID-19 vaccine uptake in the general population is very poor. In particular, to the best of our knowledge, only three studies on the factors influencing COVID-19 vaccination in the general population have been published in pre-print services (Glampson et al., 2021; McCabe et al., 2021; Nguyen et al., 2021). Furthermore, these studies investigated only certain demographic characteristics of the general population, such as gender, age, race/ethnicity educational level, profession, income, and population density. Thus, our objectives were (i) to estimate the percentage of the general population vaccinated against the COVID-19 and (ii) to investigate the factors associated with COVID-19 vaccine uptake.

## Methods

### Study design and participants

We conducted an online cross-sectional study in Greece. We included individuals over 18 years of age. By the beginning of 2021 until the time of the study, a free COVID-19 vaccine had already been offered to all adults residents of Greece by the Greek government. In more detail, on the 11th January 2021, Greece approves a COVID-19 vaccine for the general population. The Greek government initially targeted vaccination of vulnerable groups and gradually by June 2021 the ability of free vaccination was given to all adults. COVID-19 vaccination was voluntary and even those diagnosed with the COVID-19 could be vaccinated. We created an anonymous version of the study questionnaire using google forms and all the participants provided informed consent to participate in the study. The questionnaire was accompanied by a detailed explanation of the study design and was distributed through social media. The study questionnaire was available between August 10^th^ – 20^th^, 2021, and 1959 participants completed the survey during this time frame. The study protocol was approved by the Ethics Committee of the Department of Nursing, National and Kapodistrian University of Athens (reference number; 370, 02-09-2021).

### Independent variables and outcome

Independent variables included socio-demographic data of the participants and attitudes towards COVID-19 vaccination and pandemic. All survey items were self-reported. Regarding socio-demographic data, we collected information on gender, age, marital status, children <18 years old, educational level (elementary school, high school, or university degree), MSc/PhD degree, working on healthcare facilities, self-perceived financial status and health status, chronic disease, previous COVID-19 diagnosis, family/friends with previous COVID-19 diagnosis, and living with elderly people or vulnerable groups during the COVID-19 pandemic. Responses in financial status and health status ranged from 0 = “very poor” to 4 = “very good”. We measured seasonal influenza vaccination in 2020 with “yes/no” answers. Also, we measured self-perceived severity of COVID-19, self-perceived knowledge regarding COVID-19 and COVID-19 vaccines, trust in COVID-19 vaccines, trust in the government, scientists, and family doctors regarding the information about the COVID-19 vaccines, and concerns about the side effects of COVID-19 vaccination. Responses ranged from 0 to 10 with higher values indicate higher levels of self-perceived severity of COVID-19, knowledge, trust, and concerns. We averaged item responses to create continuous variables.

Our outcome variable was COVID-19 vaccination status, measured through “yes/no” answers. Also, we collected information on the possible reasons for the decline of COVID-19 vaccination.

### Statistical analysis

We used numbers (percentages) to present categorical variables and mean (standard deviation) to present continuous variables. To investigate predictors of COVID-19 vaccine uptake, we used logistic regression analysis. First, a univariate logistic regression analysis was fit to identify which variables were associated with accepting a COVID-19 vaccine. Then, a multivariate logistic regression analysis was applied to find the independent predictors of COVID-19 vaccination. The multivariate model included the variables that in the univariate analysis had a p-value <0.05. A backward stepwise model was applied and adjusted odds ratios (OR), 95% confidence intervals (CI), and two-sided p-values were presented. In multivariate model, p-value<0.05 was considered significant. Statistical analysis was performed with the Statistical Package for Social Sciences software (IBM Corp. Released 2012. IBM SPSS Statistics for Windows, Version 21.0. Armonk, NY: IBM Corp.).

## Results

Detailed socio-demographic characteristics of the 1959 participants are shown in Table 1. Mean age of the participants was 41.5 years old. Most of the participants were females (75.3%) and married (64.3%). Among them, 81% had a University degree and 42.4% had a MSc/PhD degree. Regarding the COVID-19 status, 9.5% of the participants were diagnosed with COVID-19 and 57% had family/friends with a previous COVID-19 diagnosis. The majority of the participants considered their financial status as moderate/good (83%) and their health status as good/very good (79.1%).

**Table 1.** Socio-demographic characteristics of the participants.

| Characteristics | N | % |
| --- | --- | --- |
| Gender |  |  |
| Females | 1476 | 75.3 |
| Males | 483 | 24.7 |
| Age (years)<sup>a</sup> | 41.5 | 10.6 |
| Marital status |  |  |
| Singles | 553 | 28.2 |
| Married | 1260 | 64.3 |
| Widowed | 135 | 6.9 |
| Divorced | 11 | 0.6 |
| Children <18 years old |  |  |
| No | 919 | 46.9 |
| Yes | 1040 | 53.1 |
| Educational level |  |  |
| Elementary school | 28 | 1.2 |
| High school | 345 | 17.6 |
| University degree | 1586 | 81.0 |
| MSc/PhD degree |  |  |
| No | 1129 | 57.6 |
| Yes | 830 | 42.4 |
| Working on healthcare facilities |  |  |
| No | 1036 | 52.9 |
| Yes | 923 | 47.1 |
| Self-perceived financial status |  |  |
| Very poor | 40 | 2.0 |
| Poor | 194 | 9.9 |
| Moderate | 1096 | 55.9 |
| Good | 530 | 27.1 |
| Very good | 99 | 5.1 |
| Self-perceived health status |  |  |
| Very poor | 9 | 0.5 |
| Poor | 50 | 2.6 |
| Moderate | 351 | 17.9 |
| Good | 973 | 49.7 |
| Very good | 576 | 29.4 |
| Chronic disease |  |  |
| No | 1522 | 77.7 |
| Yes | 437 | 22.3 |
| Previous COVID-19 diagnosis |  |  |
| No | 1773 | 90.5 |
| Yes | 186 | 9.5 |
| Family/friends with previous COVID-19 diagnosis |  |  |
| No | 842 | 43.0 |
| Yes | 1117 | 57.0 |
| Living with elderly people or vulnerable groups during the COVID-19 pandemic |  |  |
| No | 1354 | 69.1 |
| Yes | 605 | 30.9 |

<sup>a</sup> mean, standard deviation

We presented participants’ attitudes towards COVID-19 vaccination and pandemic in Table 2. Most participants had been vaccinated against the COVID-19 (87.8%), while about half had been vaccinated against influenza (52.5%). The main reasons for refusing COVID-19 vaccination were concerns about the safety and effectiveness of COVID-19 vaccines (43.8%), concerns about the side effects of COVID-19 vaccines (22.7%), previous COVID-19 diagnosis (10.7%), and females’ effort to get pregnant (8.2%). Self-perceived severity of COVID-19 and concerns about the side effects of COVID-19 vaccination were moderate, while knowledge regarding COVID-19 and COVID-19 vaccines was high. Participants had more confidence in family doctors and scientists than in the government regarding the information about the COVID-19 vaccines.

**Table 2.** Participants’ attitudes towards COVID-19 vaccination and pandemic.

|  | N | % |
| --- | --- | --- |
| COVID-19 vaccination |  |  |
| No | 239 | 12.2 |
| Yes | 1720 | 87.8 |
| Seasonal influenza vaccination in 2020 |  |  |
| No | 931 | 47.5 |
| Yes | 1028 | 52.5 |
| Reasons for decline of COVID-19 vaccination |  |  |
| I have doubts about the safety and effectiveness of COVID-19 vaccines | 102 | 43.8 |
| I am afraid of side effects of COVID-19 vaccines | 53 | 22.7 |
| I believe that I will not be infected by COVID-19 | 1 | 0.4 |
| I believe that even if I get infected with COVID-19, nothing bad will happen to me | 7 | 3.0 |
| I have already been diagnosed with COVID-19 and the vaccine will not be beneficial for me | 25 | 10.7 |
| I am afraid because I suffer from a chronic disease | 8 | 3.4 |
| Family physician does not allow me to take a COVID-19 vaccine due to my medical condition | 4 | 1.7 |
| My religion does not allow me to take a COVID-19 vaccine | 3 | 1.3 |
| I am trying to get pregnant | 19 | 8.2 |
| I am afraid because I am pregnant | 11 | 4.7 |
| Self-perceived severity of COVID-19<sup>a</sup> | 4.6 | 2.7 |
| Self-perceived knowledge regarding COVID-19<sup>a</sup> | 8.9 | 1.3 |
| Self-perceived knowledge regarding COVID-19 vaccines<sup>a</sup> | 8.6 | 1.7 |
| Trust in the government regarding the information about the COVID-19 vaccines<sup>a</sup> | 5.2 | 3.4 |
| Trust in scientists regarding the information about the COVID-19 vaccines<sup>a</sup> | 7.5 | 2.9 |
| Trust in family doctors regarding the information about the COVID-19 vaccines<sup>a</sup> | 8.2 | 2.1 |
| Concerns about the side effects of COVID-19 vaccination<sup>a</sup> | 5.6 | 3.1 |
| Trust in COVID-19 vaccines<sup>a</sup> | 7.3 | 2.8 |

<sup>a</sup> mean, standard deviation

Results of the univariate and multivariate logistic regression analysis are presented in Table 3. Multivariate logistic regression analysis identified that increased age was associated with an increased likelihood of COVID-19 vaccination. Also, participants with a higher level of education and those working in health services had a greater probability to take a COVID-19 vaccine. Participants without a previous COVID-19 diagnosis and those with previous seasonal influenza vaccination history were more frequently vaccinated against the COVID-19. Additionally, increased self-perceived severity of COVID-19, knowledge regarding COVID-19, and trust in COVID-19 vaccines and scientists regarding the information about the COVID-19 vaccines were associated with COVID-19 vaccine uptake. On the other hand, the likelihood of vaccination was lower for participants who were more concerned about the side effects of COVID-19 vaccination and those who trusted family doctors more for COVID-19 vaccine information.

**Table 3.**
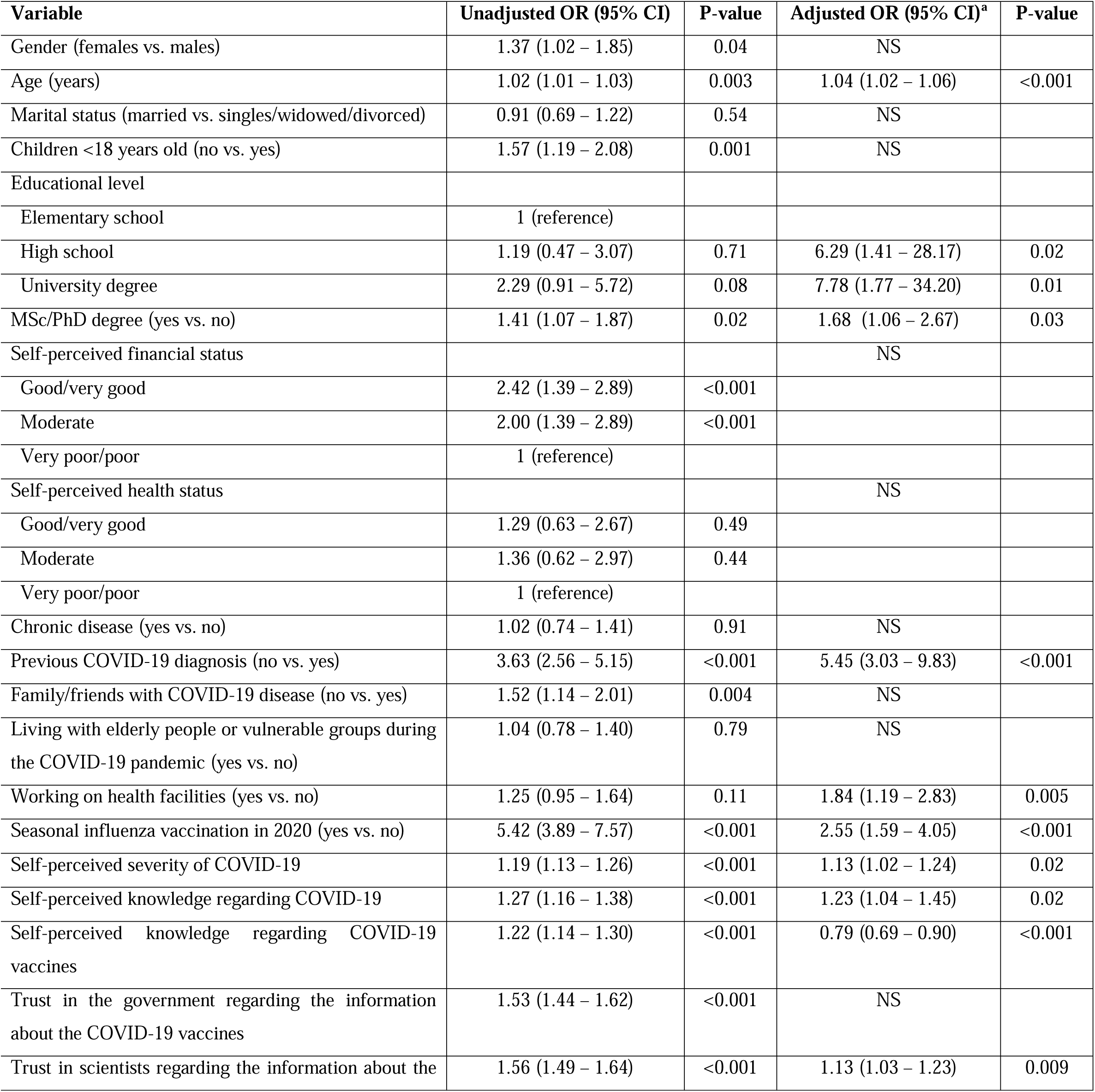
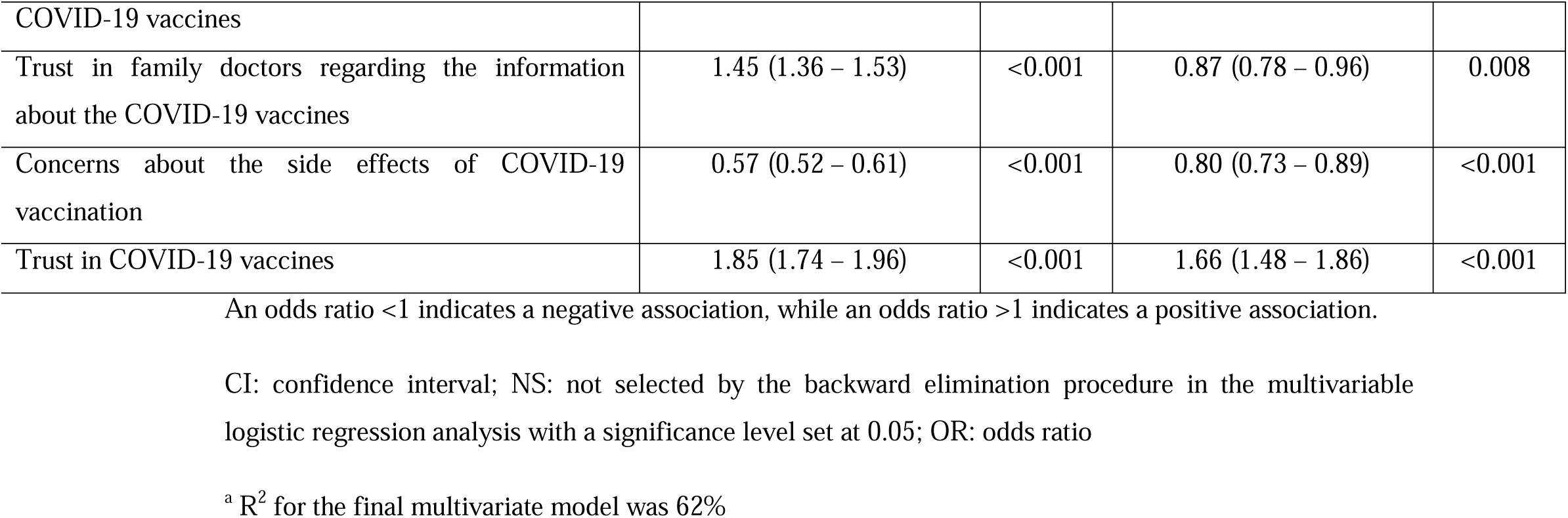
Univariate and multivariate logistic regression analysis with COVID-19 vaccine uptake among the participants as the dependent variable (reference: COVID-19 vaccine denial).

## Discussion

Our study investigated the factors that influence the decision of the general population to be vaccinated against the COVID-19. It is highly encouraging that the majority of participants had been vaccinated against the COVID-19 (87.8%). In three similar studies conducted to date, this rate was 28.6% in a study in the USA and the United Kingdom (Nguyen et al., 2021), 94.1% in a study in the United Kingdom (Glampson et al., 2021), and 98% in a study in the USA (McCabe et al., 2021). In addition, we found that the most important reasons why participants were not vaccinated were concerns about the safety, effectiveness and side effects of COVID-19 vaccines, previous COVID-19 diagnosis, and females’ effort to get pregnant. Literature confirms this finding with studies conducted not only in the general population but also in patients and health professionals (Galanis et al., 2021).

Another interesting finding of our study was the seasonal influenza vaccination rate in 2020. This rate (52.5%) was high since worldwide the influenza vaccination rate does not exceed 30% (Blank et al., 2008, 2009; Wang et al., 2018). However, it is possible that the high influenza vaccination rate in 2020 may be due to the findings of studies that show that influenza vaccination is associated with a lower risk of SARS-CoV-2 infection (R Wang et al., 2021). In any case, the high influenza vaccination rate is an extremely encouraging fact indicating the positive attitude of the participants towards vaccination.

We studied several demographic characteristics of participants and we found that age, educational level, and working in health services were associated with vaccination. More specifically, increased age was associated with an increased likelihood of vaccination. This finding is confirmed by studies in the general population and healthcare workers (Barry et al., 2021; Martin et al., 2021; McCabe et al., 2021). This result can be attributed to the fact that from the beginning of the COVID-19 pandemic, increased age has been found to be associated with worse outcomes such as admission to an intensive care unit and death (Mehraeen et al., 2020; Sepandi et al., 2020; Yanez et al., 2020). Probably, older people may confront COVID with more fear and anxiety, resulting in a higher rate of vaccination. This thought is strengthened by our findings that increased self-perceived severity of COVID-19 and the absence of COVID-19 diagnosis in the past were associated with an increased likelihood of COVID-19 vaccination. Studies with healthcare workers in the USA and the United Kingdom confirm these results (Martin et al., 2021; Pacella-LaBarbara et al., 2021). Risk perception regarding the COVID-19 is crucial in the decision of individuals to be vaccinated, since the intention to vaccinate is higher among those who consider the COVID-19 to be dangerous and life-threatening (Caserotti et al., 2021; Glöckner et al., 2020; Karlsson et al., 2021; Ward et al., 2020). In addition, it is possible that past COVID-19 patients may feel that they have acquired immunity against the virus and are not so afraid of the negative clinical outcomes (Pacella-LaBarbara et al., 2021). On the contrary, individuals without a history of COVID-19 infection may feel more vulnerable and therefore decide to vaccinate.

Our multivariate regression model revealed that participants with a higher level of education were more frequently vaccinated against the COVID-19. This finding echoes the results of research which show higher COVID-19 vaccine uptake among more educated individuals (Malesza and Bozym, 2021; Pacella-LaBarbara et al., 2021). In general, people with higher socioeconomic status are more frequently vaccinated against the COVID-19 (Malesza and Bozym, 2021; McCabe et al., 2021; Schrading et al., 2021). It is possible that a higher level of education enables people to better understand the vast amount of information about the pandemic. This is supported by our finding that increased knowledge regarding COVID-19 was related to an increased likelihood of vaccination. Better information about the COVID-19 pandemic and vaccines enables people to detect fake news. Thus, a relationship of trust is created between individuals and scientists reducing vaccine hesitancy (Dubé et al., 2013; Gust et al., 2005). Our study and the literature confirm this finding since increased trust in COVID-19 vaccines and scientists is associated with an increased likelihood of COVID-19 vaccination (Nguyen et al., 2021). Moreover, increased trust in COVID-19 vaccines and scientists could reduce concerns about the safety and effectiveness of COVID-19 vaccines that are the main reason for decline of COVID-19 vaccination (Gibbon et al., 2021; Malesza and Bozym, 2021; Nguyen et al., 2021; Schrading et al., 2021).

We found that participants with previous seasonal influenza vaccination history had a greater probability to take a COVID-19 vaccine. The relationship between influenza vaccination and the COVID-19 vaccine uptake in the general population has not been investigated but a systematic review has already shown the positive effect of influenza vaccination on the intention of healthcare workers to accept a COVID-19 vaccine (Galanis et al., 2020). Acceptance of influenza vaccination is evidence of positive attitudes towards vaccination which also increases the likelihood of COVID-19 vaccine uptake even though knowledge of the general population regarding COVID-19 vaccines is much less than that of influenza vaccines.

### Limitations

Our study had certain limitations. Our study population was large, but not representative since we used a convenience sample. For instance, the proportion of women, people with a high level of education, and workers in healthcare facilities was considerably higher in our study compared to the general population in Greece. Moreover, online surveys during the COVID-19 pandemic are a rational approach but diminish the representativeness of the study population since individuals with limited internet access are less likely to participate. Additionally, the response rate and the profile of non-correspondents cannot be estimated in online surveys. Thus, it would be wise not to generalize our conclusions but to carry out studies with more representative samples. Information bias was possible in our study since the study questionnaire was self-administered and we cannot objectively verify self-reported vaccination. Anonymity in our study may have reduced this information bias. Finally, we have investigated several factors that may affect individuals’ decision to uptake a COVID-19 vaccine, but there may be other factors influencing this decision.

## Conclusions

We observed a high level of COVID-19 vaccine uptake in the Greek general population during the first seven months of the vaccine rollout. We found that several factors influence the decision of the general population to get vaccinated against the COVID-19, such as age, educational level, previous COVID-19 diagnosis, previous seasonal influenza vaccination history, etc. Understanding the factors affecting individuals’ decision to take a COVID-19 vaccine is essential to improve the COVID-19 vaccination coverage rate. Further research with more representative samples is needed to better understand how people decide to uptake a COVID-19 vaccine. Policymakers and scientists should scale up their efforts to increase the COVID-19 vaccination rate among specific population groups such as young people, people with a low level of education, people with negative attitudes towards vaccination, etc. Optimizing protection of the general population through vaccination is crucial to control the COVID-19 pandemic.

## Data Availability

Data will be available after reasonable request

## Author Contributions

Conceptualization: Petros Galanis, Irene Vraka, Daphne Kaitelidou

Study design: Petros Galanis, Irene Vraka

Data acquisition: Petros Galanis, Ioannis Moisoglou

Formal analysis: Petros Galanis, Olympia Konstantakopoulou, Aglaia Katsiroumpa

Supervision: Petros Galanis, Daphne Kaitelidou

Writing original draft: Petros Galanis, Irene Vraka, Olga Siskou, Aglaia Katsiroumpa, Ioannis Moisoglou

Writing, review and editing: Petros Galanis, Olga Siskou, Olympia Konstantakopoulou, Daphne Kaitelidou

